# Assessing the Socio-geographic and lifestyle Factors Impacting Epithelial Ovarian Cancer Outcomes: A Retrospective Study Based on County Health Ranking in Missouri

**DOI:** 10.1101/2025.06.18.25329863

**Authors:** Carlye Goldenberg, Kavita Krell, Edgar Diaz Miranda, Sooah Ko, Maya Demirchian, Grace Anne Dyer, Mark Hunter, Erin Tuller, Amanda Hull, Lei Lei

**Affiliations:** School of Medicine, University of Missouri. Columbia, Missouri, 65201; Department of Obstetrics, Gynecology and Women’s Health, School of Medicine, University of Missouri. Columbia, Missouri, 65201

**Author notes:** Corresponding author Corresponding author: Lei Lei, Department of Obstetrics, Gynecology and Women’s Health University of Missouri School of Medicine. These authors contributed equally (Carlye Goldenberg, Kavita Krell).

## Abstract

**Objective:** This study examined how obesity, smoking, and pregnancy history, characterized as lifestyle factors, are associated with survival of epithelial ovarian cancer, and investigated whether epithelial ovarian cancer presentation, survival, and cancer recurrence are affected by patient home geographic location.

**Methods:** A retrospective analysis was conducted on all patients with epithelial ovarian cancer treated at the University of Missouri and Ellis Fischel Cancer Center between 2008 and 2023. Patient charts were reviewed for cancer history, lifestyle factors, patient status, laboratory values, and residential zip codes which were categorized using Missouri ZIP Health Rankings. Survival, cox univariate and multivariate logistic regression, and association analyses were performed.

**Results:** In this cohort, stage at diagnosis, histologic type, age at diagnosis and initial CA125 proved to be significant predictors of survival, while lifestyle factors including BMI, smoking, and pregnancy were not. Notably, patients residing in communities with the lowest zip code health rankings experienced higher rates of cancer recurrence, despite a lower overall number of cases compared to higher-ranked communities.

**Conclusion:** Although the lifestyle factors investigated in this study were not significantly associated with survival, a geographic disparity in recurrence rates and total cases was clear, suggesting possible underdiagnosis and barriers to accessing care in lower ranked zip codes. These findings emphasize an evident need to further investigate community-specific healthcare access and delivery, as well as other lifestyle factors that may be contributing to these differences.

## 1. Introduction

Ovarian cancer is the seventh most common cancer among women globally and has the highest mortality rate among gynecological tumors^1^. In 2024, an estimated 19,680 new cases of ovarian cancer were reported, along with approximately 12,740 deaths in the United States^2^. This high mortality rate can be attributed to several factors, one of which is that ovarian cancer presents with non-specific symptoms at the early stage, unlike other female cancers that have more obvious early warning signs. As a result, it is often diagnosed in advanced stages (III or IV)^3^. Therefore, identifying factors, such as lifestyle, geographic, and socioeconomic influences, that could facilitate early detection and prevention opportunities is paramount.

Per classification by the 2020 World Health Organization, most malignant ovarian cancers are epithelial cancer (90%). Germ cell (5%) and sex cord-stromal (2-5%) tumors occur with relatively low frequency^4,5^. While extensive studies have been conducted to identify lifestyle factors influencing survival, findings have been inconsistent. Some studies have reported a positive association between severe obesity and ovarian cancer risk, particularly in the premenopausal period, while others have found no significant relationship^5,6^. One study observed a reduced risk of all histologic subtypes of epithelial ovarian cancer in parous women, with the risk decreasing as parity increased^7^. Contrastingly, a large population-based study noted that parity was linked to better outcomes in germ cell tumors; however, it did not significantly affect the prognosis of epithelial ovarian cancer subtypes^8^. Nevertheless, certain risk factors, such as initial CA125, age and stage at diagnosis are established predictors of outcome, with younger women generally having higher survival rates than older women^9–11^.

Geographic location remains a relatively underexplored factor in ovarian cancer mortality despite growing evidence that where a person lives may be more predictive of their health outcomes than their genetic code^12^. Previous research regarding health outcomes in Missouri identified the rural areas of Southeastern Missouri and the City of St. Louis as having the lowest survival rates for ovarian cancer in the state^13^. The Missouri ZIP Health Rankings Project, founded by the Robert Wood Johnson Foundation, is a partnership between the Washington University School of Medicine and the Hospital Industry Data Institute that measures community health at the ZIP code level using hospital discharge and census data within the County Health Rankings model^14^. The County Health Rankings (CHR) evaluate critical health factors such as high school graduation rates, obesity, smoking, unemployment, access to healthy foods, air and water quality, income inequality, and teen birth rates^15^.

The present study aimed to identify factors influencing survival in epithelial ovarian cancer patients treated at the University of Missouri and Ellis Fischel Cancer Center to enhance early detection and prevention efforts in rural areas. The primary objective was to examine lifestyle factors—specifically obesity, history of smoking, or pregnancy, and their relationship to ovarian cancer survival while accounting for stage and age, which are established factors influencing survival. A secondary objective was to explore how geographic location affects presentation and survival, using initial CA125 and the Missouri County Health Rankings (CHR) to help identify areas where early detection and screening efforts may be most needed.

## 2. Methods

### 2.1. Patient data collection and categorization

A retrospective analysis was conducted on all patients with epithelial ovarian cancer treated at the University of Missouri and Ellis Fischel Cancer Center between 2008 and 2023 (IRB#2099034). As seen in Figure 1, patients were selected using the ICD-10 site code "C56.9 - Ovary" from the University of Missouri Health Care Cancer Registry, identifying 293 cases. The initial cohort included all patients diagnosed with ovarian cancer who had received surgery, chemotherapy, radiation therapy, hormone therapy, or immunotherapy or had chosen no treatment at the University of Missouri. ICD-10 histology codes categorized patients to identify those with epithelial ovarian cancer. Non-primary epithelial ovarian cancers were excluded. Additionally, cases with initial treatment at another hospital, without confirmation of primary epithelial ovarian cancer, or simultaneous diagnosis of multiple primary cancers were removed. The final dataset consisted of 219 patients (**Figure 1**).

**Figure 1.**
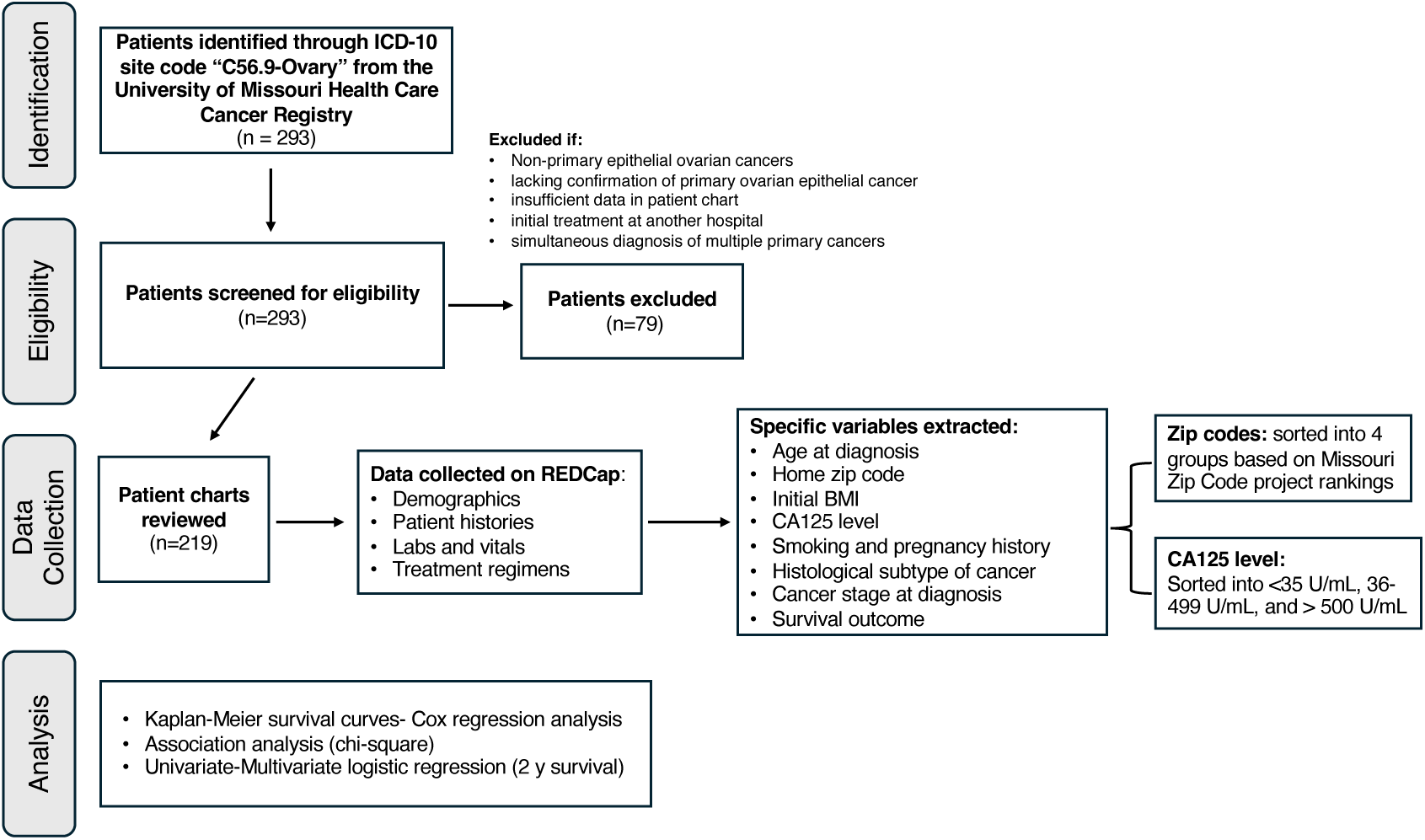
Study methodology and patient selection flowchart.

Demographic and clinical data were collected utilizing REDCap, a secure web application for building and managing online surveys and databases. Information was recorded from clinical notes with gynecologic oncology, including demographic information, patient medical and social histories, lab work, vitals, and treatment regimens. We also documented the tumor histologic subtype and stage of disease via pathology report, and recurrence via most recent gynecologic oncology note.

Patient status (alive, deceased, or unknown) was determined based on the last date of contact in the hospital records. Patients with documented contact within the last 2 years of the study period were classified as alive. Patients with no contact within this period and without a recorded death date were classified as status unknown. Patients with a confirmed death date in hospital records were classified as deceased. Patients discharged to hospice care were assumed to have died within the same year of discharge unless otherwise specified in the medical records. This assumption was based on the typical prognosis of hospice-enrolled patients.

Cancer recurrence was recorded as part of this study. Recurrence was classified as ’Yes’ if explicitly documented in the most recent gynecologic oncology note, including cases in which recurrence was not proven but it was noted by gynecologic oncology experts to be “likely recurrence.” It was classified as ’No’ if the patient was either deceased or alive with no mention of recurrence in the medical records. Recurrence was classified as ’Unknown’ if the patient’s status was unknown and there was no prior documentation of recurrence in earlier gynecologic oncology notes.

We categorized CA125 levels as level 1 (under 35 U/mL), level 2 (between 36 and 499 U/mL), and level 3 (over 500 U/mL). For CHR, information was collected from the Missouri Zip Code project using the map feature (**Supplementary Figure 1**). A map of Missouri was curated, ranking each zip code from 1 to 934, with a lower value indicating more optimal health factors. The ranking was built based on critical health factors, such as high school graduation rates, obesity, smoking, unemployment, access to healthy foods, air and water quality, income inequality, and teen birth rates. The zip codes were grouped into five categories based on rankings: group 1 (ranks 1-235), group 2 (ranks 236-470), group 3 (ranks 471-705), group 4 (ranks 706 and above), and no data. Total population and female populations for each zip code were also recorded using the same database. Zip codes were again categorized into 4 groups: <1,001 women,1,001-2,500 women, 2,501 - 1,000 women, and >10,000 women.

### 2.2. Statistical analysis

Data analysis was performed in SAS OnDemand environment. For association analysis, 219 patients were used. For survival analysis and Univariate and multivariate logistic regression analysis, five patients were excluded due to lack of values. Association between variables and stage of cancer at initial diagnosis and CHR were analyzed by Chi-square test (FREQ Procedure).

### 2.3. Imputation

Variables with a maximum of 5% missing data were imputed^16^. The continuous variables BMI (0.47% missing), CA125 (2.80% missing), and CHR (2.34% missing) were imputed by mean imputation and then converted into categorical variables Obese (BMI > 30, Yes; otherwise, No), CA125 (groups 1 to 3), and CHR (groups 1 to 4). Categorical variables Smoking (0.47% missing), Pregnancy (2.34% missing), Stage (0.47% missing), and Type (0.47% missing) were imputed using mode imputation.

Due to the risk of multicollinearity, when the categorical explanatory variables were strongly correlated (Cramer’s V > 0.20), only one of them was chosen for the analysis. Quantitative explanatory variables were evaluated by Pearson correlation, and variables were considered highly correlated when r > 0.70.

### 2.4. Survival analysis

Overall survival was analyzed by survival analysis (Lifetest Procedure) and the comparisons among the strata were performed by Log-Rank test. Patients with unknown status were entered in the analysis as censored observations. Cox’s proportional hazard analysis was performed to determine the way in which the time of survival was affected by the co-variables (Phreg Procedure). The hazard ratios were estimated with 95% confidence intervals (CI). The significance level adopted was 0.05.

### 2.5. Univariate and multivariate logistic regression

When using univariate logistic regression (Logistic Procedure) event/trial model, the event is the number of cases of ovarian cancer and number of trials is the population. The CHR rank was considered the explanatory variable. Ls-means were compared by Tukey test with significance level α = 0.05.

For multivariate logistic regression analysis (Logistic Procedure), patients with unknown and alive status, but with the last point of contact less than two years from diagnosis, were removed, leaving 189 patients in the dataset. The order of the explanatory variables in the forward selection was designed according to their prognostic value. The Akaike Information Criterion (AIC) was estimated at each stage of forward selection and used to balance the fit and parsimony of the model. If the AIC decreased, then the respective independent variable was kept in the model at the next stage.

Sequentially, explanatory variables were used to predict the probability of survival for at least 2 years by multivariate logistic regression, variables were kept in the final model when P < 0.20. Ls-means were compared by Tukey test at p < 0.05. Probability was calculated by the general formula^17^:

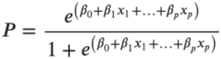

## 3. Results

### 3.1. Summarized characteristics of the study cohort

The study included 219 female patients with epithelial ovarian cancer. Most were aged 45–65 years (n=110), with 37 patients younger than 45, and 72 patients older than 65. Obesity was present in 96 patients, and 94 had a history of smoking. Pregnancy history was reported in 172 patients. The majority had advanced-stage cancer (n=135), with serous cystadenocarcinoma being the most frequent histological subtype (n=108). At data collection, 98 patients were alive, 86 had died, and 35 had unknown status (**Table 1**).

**Table 1.** Characteristics of the study cohort (n=219 patients).

| Variable | Classifications | Number of patients (%) |
| --- | --- | --- |
| Age at diagnosis (years) | Younger than 45 | 37 (16.9%) |
|  | 45 – 65 | 110 (50.2%) |
|  | Older than 65 | 72 (32.9%) |
| Obesity | No | 123 (56.2%) |
|  | Yes | 96 (43.8%) |
| History of smoking | No | 125 (57.1%) |
|  | Yes | 94 (42.9%) |
| History of pregnancy | No | 47 (21.5%) |
|  | Yes | 172 (78.5%) |
| County health ranking (CHR) | 1 | 60 (27.4%) |
|  | 2 | 92 (42.0%) |
|  | 3 | 53 (24.2%) |
|  | 4 | 14 (6.4%) |
| CA125 level | 1 ($\leq 35$ U/ml) | 41 (18.7%) |
|  | 2 (36–499 U/ml) | 97 (44.3%) |
| | 3 ($\geq 500$ U/ml) | 81 (37.0%) |
| Stage at diagnosis | Advanced | 135 (61.6%) |
|  | Early | 84 (38.4%) |
| Histological subtype of cancer | Clear cell cystadenocarcinoma (CCC) | 20 (9.1%) |
|  | Unspecified cystadenocarcinoma (UC) | 19 (8.7%) |
|  | Endometrioid adenocarcinoma (EA) | 30 (13.7%) |
|  | Mixed cell adenocarcinoma (MCA) | 19 (8.7%) |
|  | Mucinous cystadenocarcinoma (MC) | 23 (10.5%) |
|  | Serous cystadenocarcinoma (SC) | 108 (49.3%) |
| Survival status | Alive | 98 (44.7%) |
|  | Dead | 86 (39.3%) |
|  | Unknown | 35 (16.0%) |

### 3.2. Factors influencing ovarian cancer survival outcome

The Kaplan-Meier curve shown in Figure 2 illustrates the probability of survival over time in our cohort, showing a decreased survival probability with increased time (days) post diagnosis. The average time of survival was 2930.9 ± 156.8 days, with a median of 3366 days (**Supplementary Figure 2**). We then found the strongest predictors of survival utilizing Cox’s hazard analysis. The stepwise selection indicated stage at diagnosis, histological subtype of cancer, age at diagnosis, and initial CA125 level as the most influential variables (**Table 2**).

**Figure 2.**
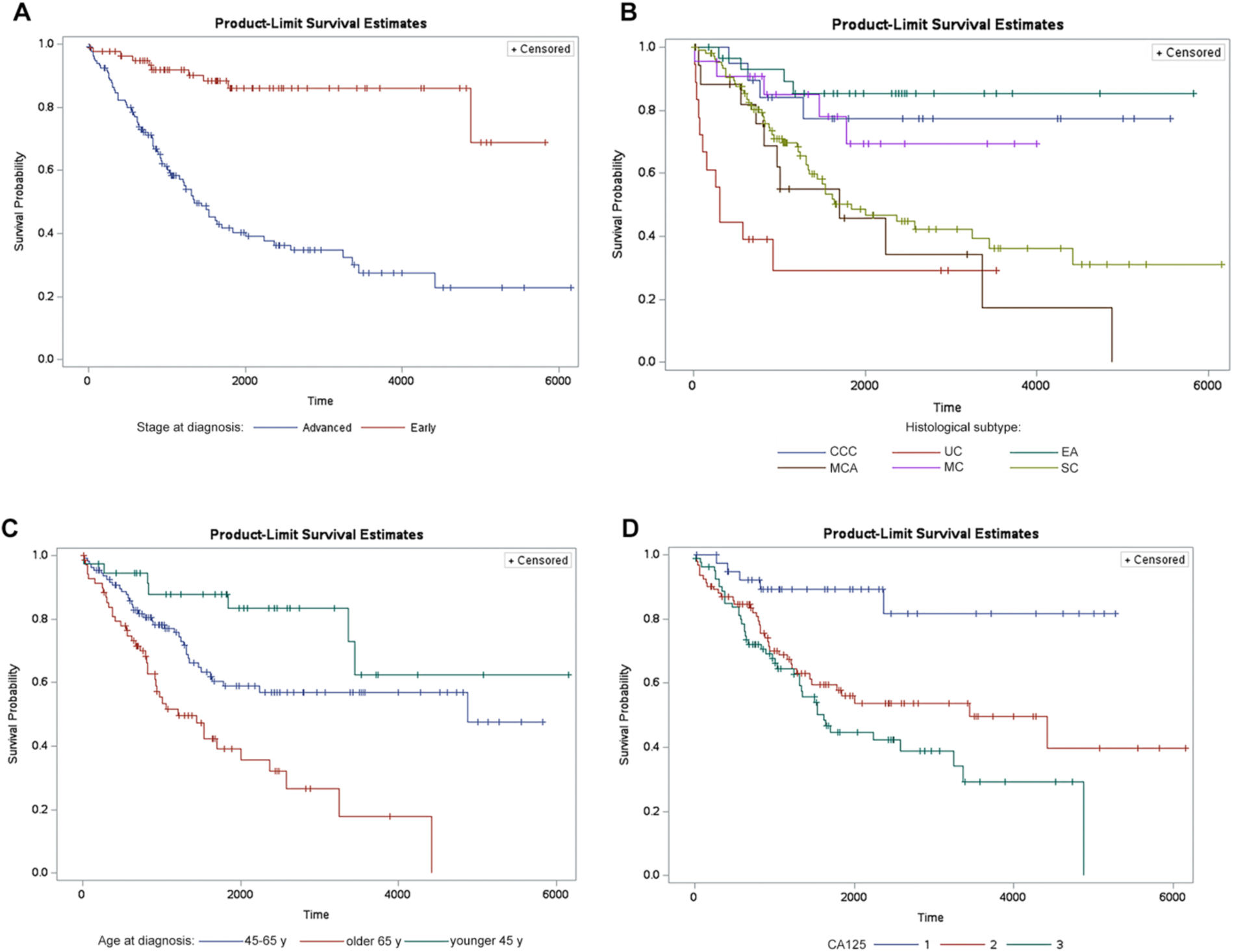
Survival curves along time (in days) based on stage at diagnosis (p < 0.0001) (**A**), histological subtype (p < 0.0001) (**B**), age at diagnosis (p < 0.0001)(**C**), and CA125 level (p = 0.0003)(**D**). Statistical difference was calculated by Log-Rank test.

**Table 2.** Summary of stepwise selection.

| Effect | DF | Summary of Stepwise Selection |  | Type 3 Tests (Final Model) |  |
| --- | --- | --- | --- | --- | --- |
|  |  | Score Chi-Square | P-value<sup>1</sup> | Wald Chi-Square | P-Value<sup>2</sup> |
| **Stage at diagnosis** | 1 | 38.2272 | <.0001 | 12.2719 | 0.0005 |
| **Histological subtype** | 5 | 19.0677 | 0.0019 | 16.7491 | 0.0050 |
| **Age at diagnosis** | 2 | 9.4590 | 0.0088 | 8.4193 | 0.0149 |
| **CA125 level** | 2 | 6.6127 | 0.0367 | 6.1694 | 0.0457 |

<sup>1</sup> Significance of variables as they were added to the model. <sup>2</sup> Final significance test for each variable in the final model.

Survival outcomes across patient groups based on those four key factors (stage at diagnosis, histological subtype, age at diagnosis, and CA125 level) were plotted and shown in **Figure 2** and **Supplementary Table 1**. Patients with advanced-stage ovarian cancer experienced a sharp decline in survival probability compared to those diagnosed at an early stage (**Figure 2A**). Survival probability declined more gradually for clear cell cystadenocarcinoma, endometrioid adenocarcinoma, and mucinous cystadenocarcinoma subtypes, whereas the decrease was more pronounced for unspecified cystadenocarcinoma, mixed cell adenocarcinoma, and serous cystadenocarcinoma subtypes (**Figure 2B**). Patients over 65 years experienced a notably steeper decline in survival probability, while those under 45 had the slowest decline; patients aged 45 to 65 exhibited an intermediate pattern (**Figure 2C**). Additionally, the survival curve for CA125 level 1 showed a more gradual decline compared to levels 2 and 3 (**Figure 2D**).

Hazard ratio analysis results in Supplementary Table 2 revealed that early-stage cancer has more than three times the chance of survival compared to advanced-stage disease. Regarding histological subtype, clear cell and endometrioid tumors were associated with better survival outcomes, whereas unspecified cystadenocarcinoma and mixed cell adenocarcinoma had the poorest prognoses. Patients under 45 years old demonstrated higher survival rates than older age groups (**Supplementary Table 2**).

### 3.3. Associations between the stage at diagnosis and health factors

We investigated whether the stage at diagnosis (early vs. advanced) was associated with various lifestyle and health factors (**Figure 3**). Our analysis showed that an advanced-stage diagnosis was significantly linked to older age, cancer recurrence, elevated CA125 level at the initial visit, and specific histological subtypes (**Figure 3A, F, G, H**). Notably, 87.4% of patients diagnosed at an advanced stage were over 45 years old. Cancer recurrence occurred in 42.2% of advanced-stage cases, compared to only 14.3% in early-stage cases. 48.9% of patients with advanced disease had an initial CA125 level above 500 U/ml, whereas only 17.9% of early-stage cases did. 65% of serous cystadenocarcinoma cases were diagnosed at an advanced stage. However, the stage at diagnosis was not significantly associated with obesity, smoking history, pregnancy history, or CHR score (**Figure 3B-E**).

**Figure 3.**
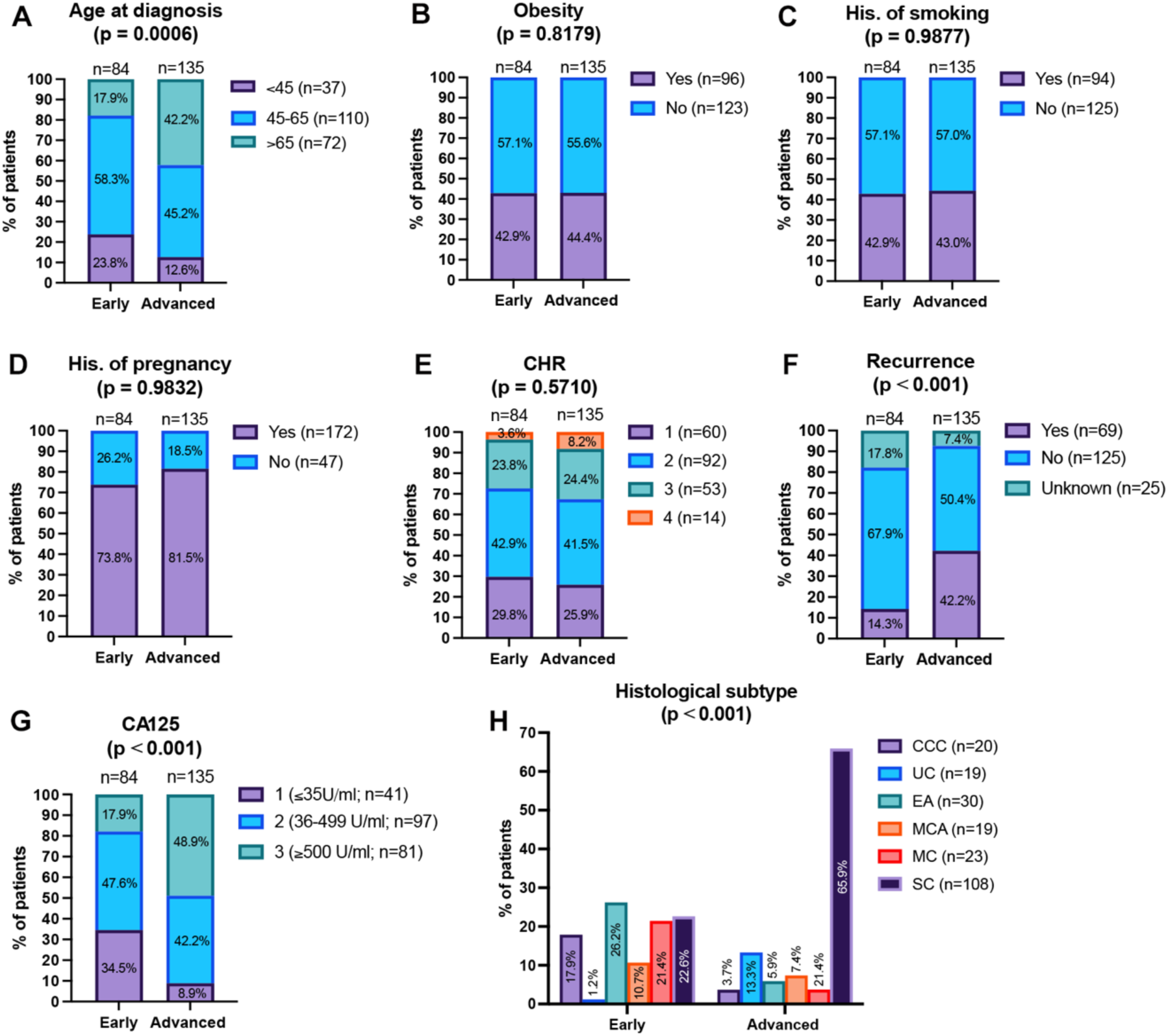
Association analysis between stage at diagnosis (early or advanced) and age at diagnosis (**A**), obesity (**B**), history of smoking (**C**), history of pregnancy (**D**), CHR (**E**), recurrence (**F**), CA125 level (**G**), and histological subtype of cancer (**H**): clear cell cystadenocarcinoma (CCC), unspecified cystadenocarcinoma (UC), endometrioid adenocarcinoma (EA), mixed cell adenocarcinoma (MCA), mucinous cystadenocarcinoma (MC), serous cystadenocarcinoma (SC).

### 3.4. Associations between CHR and health factors and ovarian cancer diagnosis

We examined whether the CHR score was associated with health factors, including age at diagnosis, obesity, smoking history, and pregnancy (**Figure 4A-D**). No significant association was observed, although patients in CHR Group 4 were statistically more likely to have a history of smoking.

**Figure 4.**
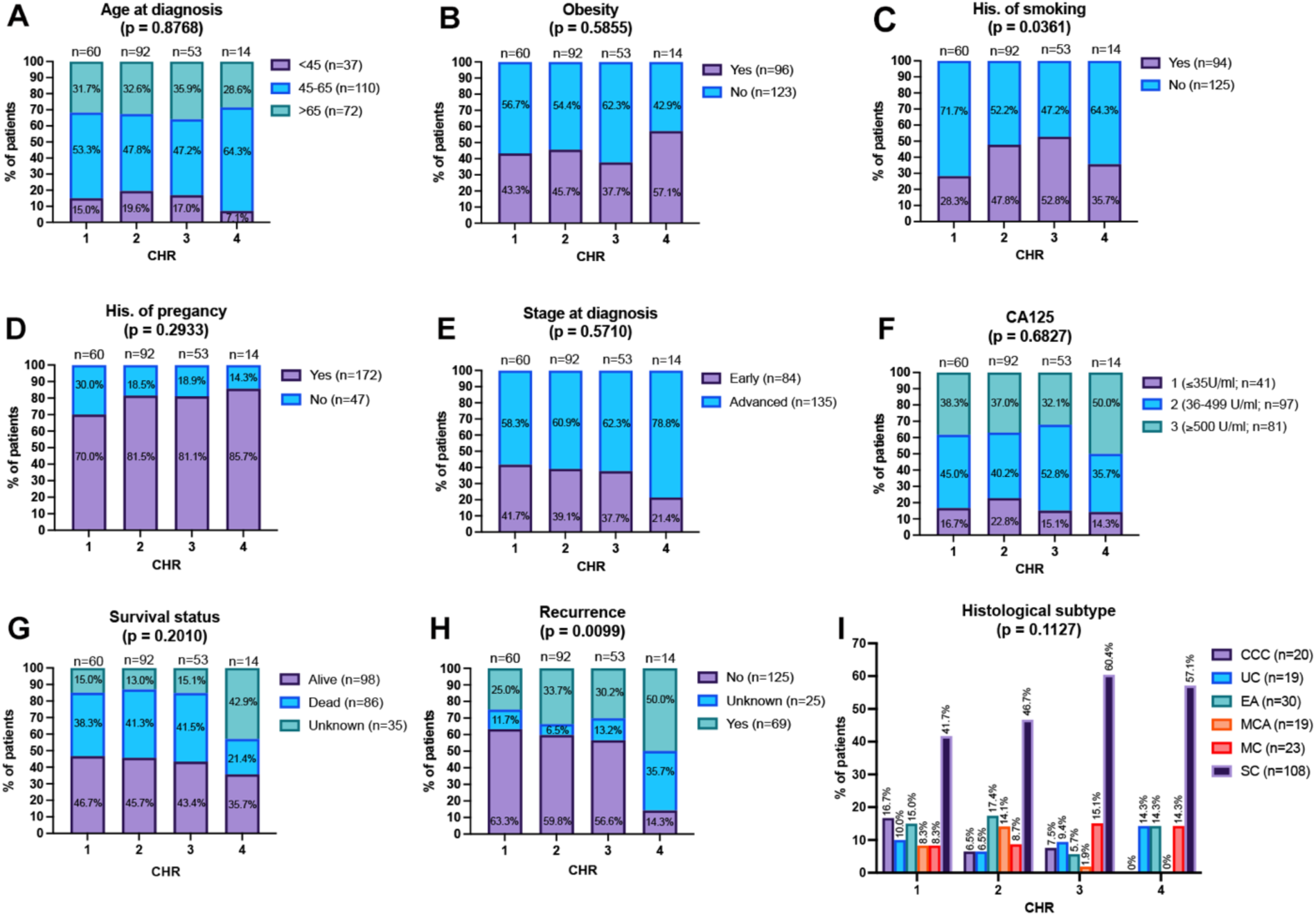
Association analysis between CHR score and age at diagnosis (**A**), obesity (**B**), history of smoking (**C**), history of pregnancy (**D**), stage at diagnosis (**E**), CA 125 level (**F**), survival status (**G**), recurrence (**H**), and histological subtype of cancer (**I**): clear cell cystadenocarcinoma (CCC), unspecified cystadenocarcinoma (UC), endometrioid adenocarcinoma (EA), mixed cell adenocarcinoma (MCA), mucinous cystadenocarcinoma (MC), serous cystadenocarcinoma (SC).

We further examined whether the CHR score was associated with stage at diagnosis, CA125 level, histological subtype, survival status, or cancer recurrence in our dataset (**Figure 4E-I**). No significant associations were found between CHR ranking and stage at diagnosis, CA125 level, survival status, or histological subtype.

However, our analysis revealed a significant correlation between CHR score and cancer recurrence, indicating that individuals in CHR Group 4 (the lowest ranking zip codes) had a higher likelihood of recurrence (**Figure 4H**). Furthermore, there is a strong inverse relationship between CHR rank and ovarian cancer diagnosis: as the CHR ranking worsens, the likelihood of being diagnosed with epithelial ovarian cancer at our institution decreases (**Supplementary Table 3**). For example, the odds of a patient with ovarian cancer existing (being diagnosed) in CHR Group 1 are seven-times higher than in Group 3. Notably, CHR Group 4 had significantly fewer cases per 1,000 women than the better ranking groups (**Supplementary Figure 3**).

### 3.5. The effect of health factors on two-year survival outcome

The effect of the variables on the probability of survival until two years after diagnosis was evaluated by a multivariate logistic regression. Based on AIC and p-values, cancer stage, histological subtype of cancer, and age group were selected for subsequent analysis (**Supplementary Table 4**). In the final model, only stage of cancer and histological subtype presented significant effect (p < 0.05) on the probability of survival until two years (**Supplementary Table 5**). Based on the probability means, it is possible to calculate the odds ratio; thus, it was noted that early-stage tumors presented 5.2 times higher chance of survival until two years than advanced-stage tumors (**Supplementary Table 6**). Moreover, serous cystadenocarcinoma showed 7.1 times higher chance of survival than unspecified cystadenocarcinoma tumors.

## 4. Discussion

This retrospective analysis of patients with primary epithelial ovarian cancer treated at the University of Missouri and Ellis Fischel Cancer Center from 2008 to 2023 examined the relationship between lifestyle health factors and geographic location on ovarian cancer diagnosis and survival outcomes. Our analysis found that cancer stage at diagnosis, histological subtype, age group, and initial CA125 level were significant predictors of survival. In contrast, lifestyle factors such as BMI, smoking history, and pregnancy history did not show a statistically significant association with survival outcomes. These findings are consistent with previous research and underscore the critical importance of early detection in improving prognosis for ovarian cancer patients.

We also observed that advanced-stage diagnoses were more common among older patients and those presenting with higher CA125 levels, consistent with findings in previous studies^18,19^. Additionally, advanced-stage disease was more frequently associated with certain histologic subtypes, particularly serous carcinoma, and was linked to increased rates of recurrence. These trends highlight the aggressive nature of advanced-stage ovarian cancer and the significant challenges of long-term disease management. They further emphasize the need for enhanced early detection strategies, especially in older populations who may be at greater risk for delayed diagnosis and worse outcomes.

In addition to clinical predictors, our analysis explored the association between CHR and diagnosis and recurrence. We found that patients residing in CHR Group 4, the lowest-ranked zip codes, had a significantly higher likelihood of cancer recurrence. Paradoxically, the total number of ovarian cancer cases from these lower-ranked areas was markedly lower than in higher-ranked CHR groups. Therefore, a strong inverse relationship emerged between CHR ranking and the likelihood of diagnosis at our institution: as CHR rank worsened, the odds of being diagnosed with epithelial ovarian cancer decreased. These findings suggest that women in lower-ranked health areas may face barriers to accessing healthcare services, leading to underdiagnosis and delayed presentation. The higher recurrence rates in these same communities may reflect gaps in follow-up care, further underscoring the importance of improving healthcare access and continuity in underserved populations.

Despite these important insights, several limitations should be acknowledged. First, this study was conducted at two hospitals located in Columbia, Missouri, which limits the size, racial diversity, and geographic representativeness of the sample. Additionally, the Missouri Zip Code Project, used to evaluate geographic health rankings, is currently limited to the state of Missouri. As a result, the generalizability of geographic findings to other states or regions is restricted.

The retrospective nature of the study also introduces limitations. Chart reviews, while valuable, may not fully capture patients’ lifestyle histories, as time constraints for providers and limitations in electronic medical records can lead to incomplete or inaccurate documentation. Moreover, self-reported behaviors such as smoking may be underreported due to perceived irrelevance or stigma. Finally, survival status and recurrence were determined at the time of chart review, meaning outcomes for patients diagnosed later in the study period may not reflect their complete disease course.

Given these limitations, future research is essential. Expanding the study to include multiple institutions across Missouri, the Midwest, or the broader United States would enhance the geographic and demographic diversity of the sample. This would allow for a more comprehensive evaluation of how lifestyle and geographic factors affect ovarian cancer outcomes and help determine whether the trends observed in Missouri are consistent across other regions with different healthcare infrastructures and socioeconomic profiles.

While our findings reaffirm the significant role of early detection in improving survival, future work should focus on the development of practical, cost-effective, and accessible screening tools. Because early-stage detection and younger age at diagnosis were associated with better outcomes, efforts to identify reliable early biomarkers or advanced imaging techniques are critical. Incorporating lifestyle and geographic data into personalized risk prediction models could also improve screening and surveillance efforts, particularly among high-risk and underserved populations.

## 5. Conclusion

In this study of patients with epithelial ovarian cancer treated at the University of Missouri, cancer stage at diagnosis, histologic subtype, age at diagnosis, and initial CA125 level were significant predictors of survival. In contrast, lifestyle factors such as BMI, smoking, and pregnancy history were not associated with survival outcomes. Patients residing in the lowest-ranked zip codes for community health had fewer total cases and a decreased likelihood of being diagnosed at our institution yet were more likely to experience cancer recurrence. Advanced-stage disease was also linked to higher recurrence rates, underscoring the need for improved early detection strategies and better access to care. Although lifestyle factors did not appear to influence survival in this cohort, the substantial differences in case volume and recurrence between geographic regions highlight the need for further investigation into the underlying causes of these disparities.

## Data Availability

All data produced in the present work are contained in the manuscript

## Research support

This work was supported by the University of Missouri startup fund. Dr. Diaz Miranda was supported by the Lalor Foundation. Dr. Lei was supported by R01GM126028.

## CrediT author statement

**Carlye Goldenberg:** Conceptualization, Methodology, Investigation, Data Curation, Writing-Original Draft, Writing-Review & Editing.

**Kavita Krell:** Conceptualization, Methodology, Investigation, Data Curation, Writing-Original Draft, Writing-Review & Editing.

**Edgar Diaz Miranda:** Data Curation, Visualization, Formal analysis, Writing-Review & Editing.

**Sooah Ko:** Conceptualization, Methodology, Investigation, Writing-Review & Editing.

**Maya Demirchian:** Conceptualization, Methodology, Investigation, Writing-Review & Editing.

**Grace Anne Dyer:** Visualization, Writing-Review & Editing.

**Mark Hunter:** Methodology, Resources, Writing-Review & Editing.

**Erin Tuller:** Methodology, Writing-Review & Editing.

**Amanda Hull:** Methodology, Writing-Review & Editing.

**Lei Lei:** Conceptualization, Methodology, Writing-Review & Editing, Visualization, Supervision, Project administration, Funding acquisition

---

**Supplementary Figure 1.**
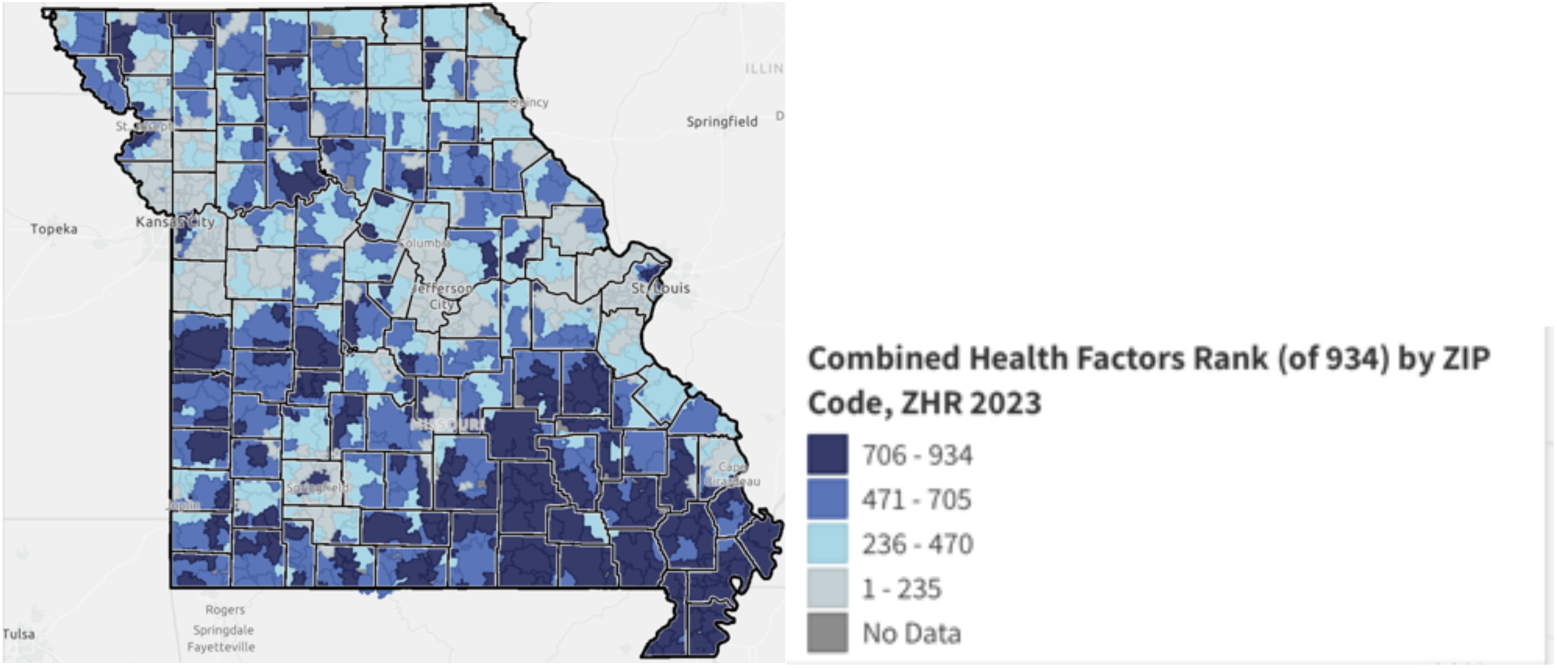
Map of Combined Health Factors Ranking by Zip Code in Missouri.

**Supplementary Figure 2.**
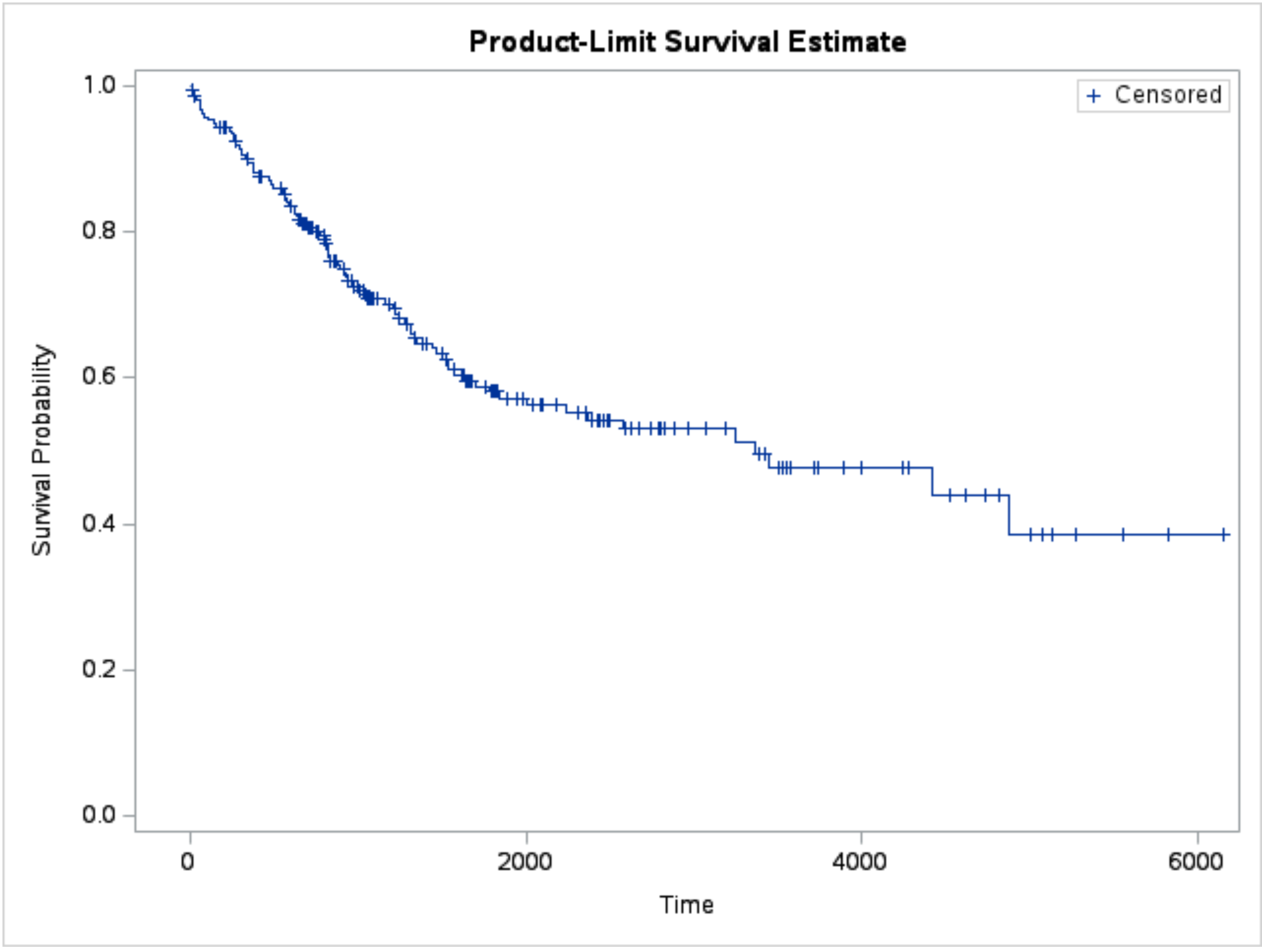
Overall survival probability along Time (in days).

**Supplementary Figure 3.**
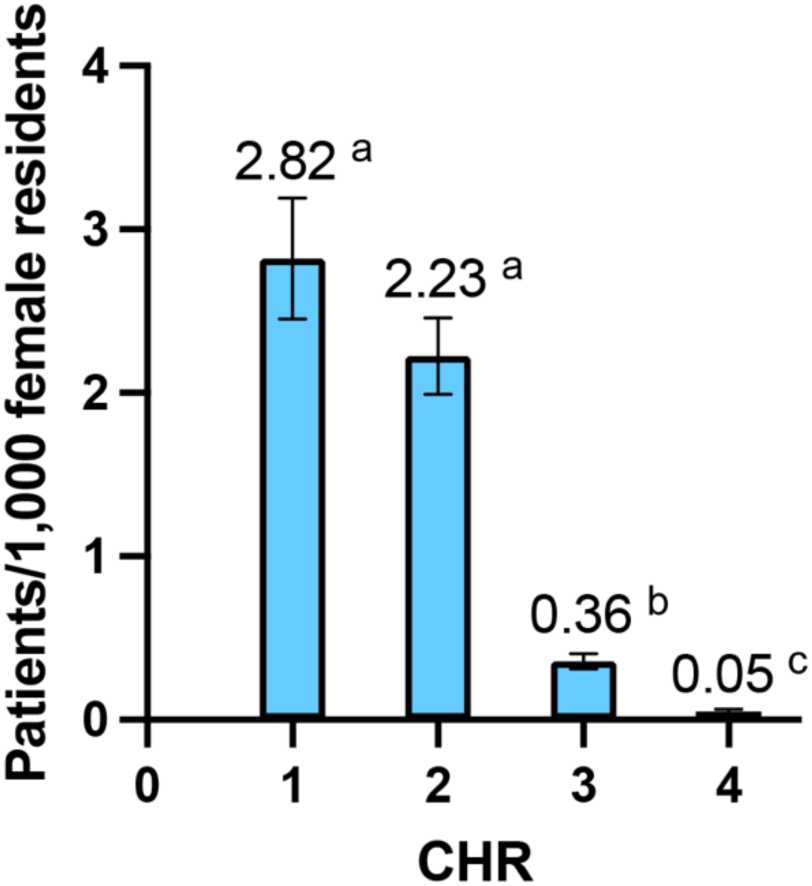
Number of cases per 1,000 women according to CHR rank. Different letters indicate difference by Tukey test (p < 0.05). The error bar presents standard error. Different letters indicate significant difference.

**Supplementary Table 1.**
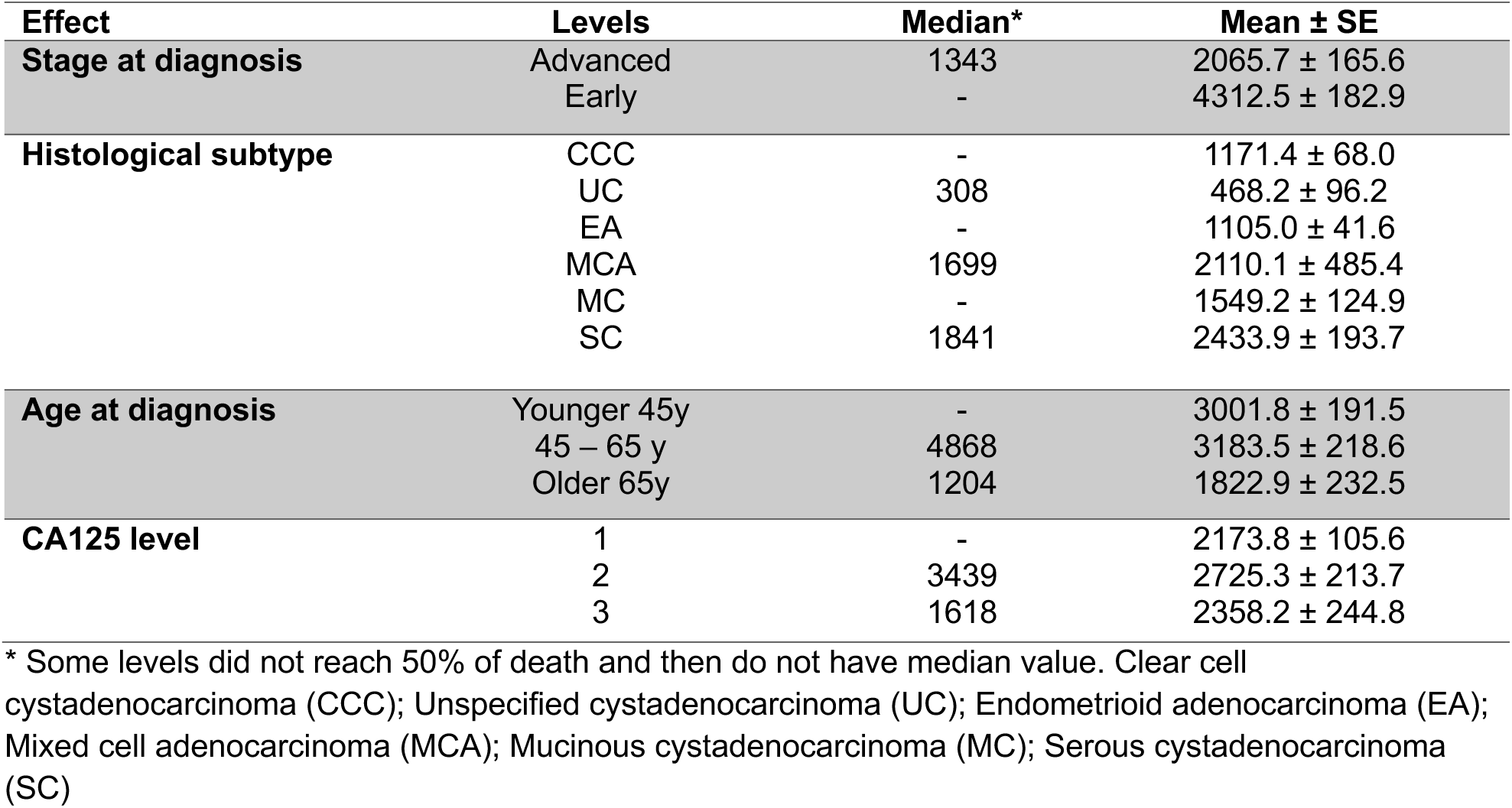
Survival time (in days) according to stage at diagnosis, cancer histological subtype, age at diagnosis, and CA125 level.

**Supplementary Table 2.** Hazard ratios and 95% confidence limits for age at diagnosis, CA125 level, stage at diagnosis, and histological subtype of cancer.

| Description | Point Estimate | 95% CL |  |
| --- | --- | --- | --- |
|  |  | Lower | Upper |
| Age at diagnosis |  |  |  |
| 45-65y vs. Older 65y | 0.755 | 0.470 | 1.213 |
| 45-65y vs. Younger 45y | 2.615 | 1.127 | 6.069 |
| Older 65y vs. Younger 45y | 3.464 | 1.488 | 8.067 |
| CA125 level |  |  |  |
| 1 vs. 2 | 0.305 | 0.116 | 0.800 |
| 1 vs. 3 | 0.416 | 0.160 | 1.084 |
| 2 vs. 3 | 1.365 | 0.831 | 2.242 |
| Stage at diagnosis |  |  |  |
| Advanced vs. Early | 3.688 | 1.777 | 7.654 |
| Histological subtype |  |  |  |
| CCC vs. UC | 0.211 | 0.063 | 0.714 |
| CCC vs. EA | 1.390 | 0.343 | 5.628 |
| CCC vs. MCA | 0.252 | 0.073 | 0.863 |
| CCC vs. MC | 0.500 | 0.130 | 1.915 |
| CCC vs. SC | 0.540 | 0.184 | 1.585 |
| UC vs. EA | 6.576 | 1.985 | 21.783 |
| UC vs. MCA | 1.190 | 0.513 | 2.761 |
| UC vs. MC | 2.364 | 0.740 | 7.550 |
| UC vs. SC | 2.554 | 1.337 | 4.881 |
| EA vs. MCA | 0.181 | 0.054 | 0.608 |
| EA vs. MC | 0.360 | 0.095 | 1.362 |
| EA vs. SC | 0.388 | 0.134 | 1.128 |
| MCA vs. MC | 1.986 | 0.624 | 6.317 |
| MCA vs. SC | 2.146 | 1.085 | 4.244 |
| MC vs. SC | 1.080 | 0.394 | 2.962 |

Clear cell cystadenocarcinoma (CCC); Unspecified cystadenocarcinoma (UC); Endometrioid adenocarcinoma (EA); Mixed cell adenocarcinoma (MCA); Mucinous cystadenocarcinoma (MC); Serous cystadenocarcinoma (SC)

**Supplementary Table 3.**
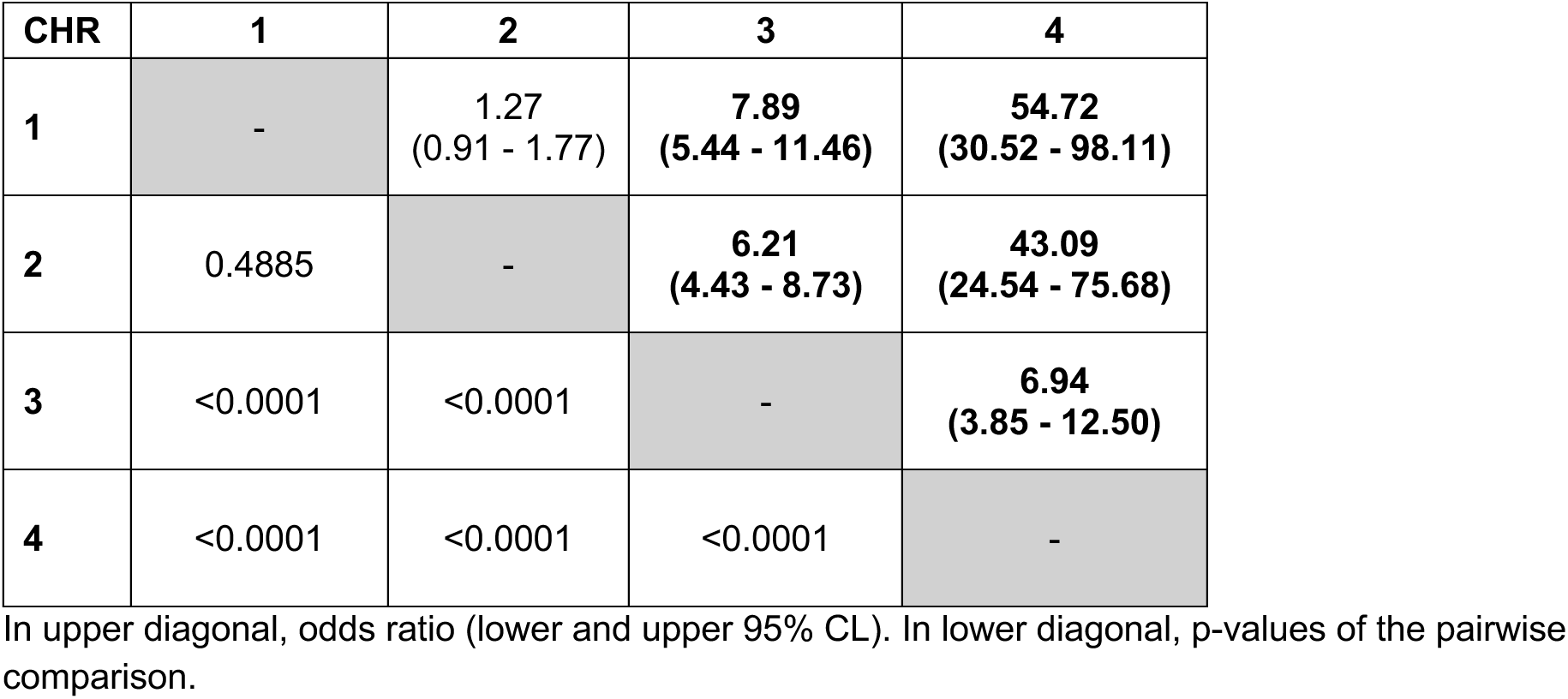
Odds of an ovarian cancer patient in the area of four CHR ranks.

| CHR | 1 | 2 | 3 | 4 |
| --- | --- | --- | --- | --- |
| 1 | - | 1.27<br>(0.91 - 1.77) | 7.89<br>(5.44 - 11.46) | 54.72<br>(30.52 - 98.11) |
| 2 | 0.4885 | - | 6.21<br>(4.43 - 8.73) | 43.09<br>(24.54 - 75.68) |
| 3 | <0.0001 | <0.0001 | - | 6.94<br>(3.85 - 12.50) |
| 4 | <0.0001 | <0.0001 | <0.0001 | - |

In upper diagonal, odds ratio (lower and upper 95% CL). In lower diagonal, p-values of the pairwise comparison.

**Supplementary Table 4.** Comparison of the Akaike’s information criterion values (AIC) from the forward selection of the explanatory variables for the logistic model and the contribution of each variable to the model fitting for dependent variable Survival until two years.

| Variable | AIC | Difference related to the "Intercept only" model | Difference related to the previous model<sup>1</sup> | p-value |
| --- | --- | --- | --- | --- |
| Intercept only | 199.693 | - | - | - |
| Stage at diagnosis | 182.186 | -17.507 | -17.507 | <0.0001 |
| Histological subtype | 178.603 | -21.090 | -3.583 | 0.0115 |
| Age at diagnosis | 178.368 | -21.325 | -0.235 | 0.1491 |
| CHR | 177.293 | -22.400 | -1.075 | 0.2083 |
| Obesity | 178.707 | -20.986 | +1.414 | 0.4430 |
| CA125 | 181.660 | -18.033 | +2.953 | 0.6067 |
| His. of pregnancy | 183.567 | -16.126 | +1.907 | 0.7622 |
| His. of smoking | 185.493 | -14.200 | +1.926 | 0.7869 |

<sup>1</sup> In forward selection, variables are added to the logistic model in decreasing order (starting with "Intercept only" model and finishing with adding "Smoking"). In each step, the AIC after adding a variable is compared to the AIC of the previous step.

**Supplementary Table 5.** Type 3 analysis of effects and analysis of maximum likelihood estimates of the multivariate multinominal logistic regression for the probability of survival until two years (Probabilities adjusted for Survival = “Yes”).

| *Type 3 Analysis of Effects* |  |  |  |
| --- | --- | --- | --- |
| Effect | DF | Wald Chi-Square | p-value |
| Age at diagnosis | 2 | 3.5161 | 0.1724 |
| Stage at diagnosis | 1 | 6.7962 | 0.0091 |
| Histological subtype | 5 | 11.1907 | 0.0477 |
| *Analysis of Maximum Likelihood Estimates* |  |  |  |
| | Estimate ± SE | Wald Chi-Square | p-value |
| Intercept | 1.2708 ± 0.8225 | 2.3869 | 0.1224 |
| Age 45-65y | 0.4084 ± 0.4199 | 0.9459 | 0.3308 |
| Age Younger 45y | 1.5571 ± 0.8559 | 3.3096 | 0.0689 |
| Age Older 65y | 0 | . | . |
| Stage Early | 1.6556 ± 0.6351 | 6.7962 | 0.0091 |
| Stage Advanced | 0 | . | . |
| Subtype CCC | -0.5974 ± 1.1003 | 0.2948 | 0.5871 |
| Subtype UC | -2.4434 ± 0.9708 | 6.3349 | 0.0118 |
| Subtype MCA | -1.0384 ± 0.9931 | 1.0933 | 0.2957 |
| Subtype MC | -1.0410 ± 1.1302 | 0.8484 | 0.3570 |
| Subtype SC | -0.4757 ± 0.8229 | 0.3342 | 0.5632 |
| Subtype EA | 0 | . | . |

Clear cell cystadenocarcinoma (CCC); Unspecified cystadenocarcinoma (UC); Endometrioid adenocarcinoma (EA); Mixed cell adenocarcinoma (MCA); Mucinous cystadenocarcinoma (MC); Serous cystadenocarcinoma (SC)

**Supplementary Table 6.** Probability estimates (LS-means) for probability of survival until two years according to age at diagnosis, cancer histological subtype, and stage at diagnosis.

| Level | Mean | Standard error | Lower mean | Upper mean |
| --- | --- | --- | --- | --- |
| *Age at diagnosis* |  |  |  |  |
| Younger 45y | 0.9384 <sup>a</sup> | 0.0460 | 0.7619 | 0.9864 |
| 45-65 y | 0.8284 <sup>a</sup> | 0.0504 | 0.7068 | 0.9062 |
| Older 65y | 0.7624 <sup>a</sup> | 0.0739 | 0.5905 | 0.8771 |
| *Histological subtype* |  |  |  |  |
| CCC | 0.8963 <sup>ab</sup> | 0.0761 | 0.6347 | 0.9772 |
| UC | 0.5770 <sup>b</sup> | 0.1695 | 0.2591 | 0.8417 |
| MCA | 0.8475 <sup>ab</sup> | 0.0885 | 0.5923 | 0.9551 |
| MC | 0.8472 <sup>ab</sup> | 0.1004 | 0.5480 | 0.9621 |
| SC | 0.9070 <sup>a</sup> | 0.0386 | 0.7992 | 0.9599 |
| EA | 0.9401 <sup>ab</sup> | 0.0456 | 0.7622 | 0.9872 |
| *Stage at diagnosis* |  |  |  |  |
| Early | 0.9339 <sup>a</sup> | 0.0355 | 0.8206 | 0.9776 |
| Advanced | 0.7297 <sup>b</sup> | 0.0712 | 0.5709 | 0.8456 |

Different letters indicate difference by Tukey test ( $p < 0.05$ ). Clear cell cystadenocarcinoma (CCC); Unspecified cystadenocarcinoma (UC); Endometrioid adenocarcinoma (EA); Mixed cell adenocarcinoma (MCA); Mucinous cystadenocarcinoma (MC); Serous cystadenocarcinoma (SC)

